# Cognitive impairment is a risk factor for long-term outcomes in septic patients: A retrospective observational study

**DOI:** 10.1101/2024.01.04.24300855

**Authors:** Hiroyuki Koami, Yuichiro Sakamoto, Yutaro Furukawa, Kosuke Mouri, Ayaka Matsuoka, Kota Shinada, Kento Nakayama, Sachiko Iwanaga, Shogo Narumi, Mayuko Koba

**Author notes:** Corresponding author: (HK).

## Abstract

The importance of long-term outcomes after sepsis has been increasingly emphasized in recent years; however, little is known about the relationship between pre-sepsis patient characteristics and long-term outcomes. This study examined the impact of patient characteristics pre-admission, specifically physical, cognitive, and emotional disorders, on long-term clinical outcomes of sepsis. From August 2014 to July 2016, this study included adult patients admitted to our hospital with a diagnosis of sepsis who were followed up for one year. These 76 patients were classified as Survivors (n=31) or Deceased (n=45), based on their one-year outcomes. In this single-center, retrospective study, 59% of sepsis patients died within one year. Multiple logistic regression models employing significant parameters from univariate analysis revealed that cognitive impairment (CI) is an independent risk factor for 1-year outcome [OR 0.184, 95% CI 0.047-0.722, P=0.0152]. Furthermore, through another univariate analysis and a log-rank test between the CI group and the Control group, created by reclassifying the same 76 patients, CI was significantly associated with a lower 1- year survival rate (25.0% vs 50.0%, P=0.03, log-rank P=0.0332). The composite outcome of either death or being bedridden at the point of discharge was also significantly higher in the CI group than the Control group (100.0% vs 75.0%, P=0.0028). These results indicate that pre-septic CI is a reliable, independent predictor of poor long-term clinical outcomes for sepsis patients.

## Introduction

Sepsis is a pressing societal issue with a global impact [1]. The Surviving Sepsis Campaign Guidelines (SSCG), launched in 2004, have significantly enhanced awareness of sepsis and have improved the quality of sepsis care [2]. Recent estimates suggest that 31.5 million cases of sepsis occur globally each year, including 19.4 million cases of severe sepsis and 5.3 million possible deaths, primarily in high-income countries[1].

Although numerous publications indicate that advances in multidisciplinary sepsis treatment have increased survival rates, sepsis constitutes an ongoing public health issue, rather than just an acute, severe illness [3, 4]. Among sepsis patients who survive until hospital discharge, as many as one-third will be dead within a year [5, 6]. Aggregated reports of observational studies documenting multiple endpoints reveal 1-year mortality rates ranging from 21.5-71.9% [7]. Moreover, deaths from sepsis continue to be reported, even several years after discharge from the hospital [8]. Given the above, the validity of 28-day mortality as an endpoint for clinical sepsis studies seems unwise [7].

As for long-term prognosis, not only death from sepsis, but also persistent disabilities such as physical impairment, cognitive impairment, and mental health problems are challenging [5]. Elderly survivors of critical care have a high risk of developing dementia within three years after discharge from the ICU [9]. Septic events can be a turning point for older patients, causing functional and cognitive impairments that impact independent daily living. This can result in additional functional limitations, re-admission to the ICU, and poor outcomes [10]. Several recent studies suggest that cognitive impairment increases by 16.7 to 33.3% during hospital stays following sepsis [10–12]. In addition, patients with cognitive impairment and ADL dependence have shorter survival rates [13].

Despite the wealth of data on post-septic cognitive impairment and its association with clinical outcomes, there are few reports regarding the correlation between pre-septic cognitive impairment and long-term outcomes. In the present study, we retrospectively collected comprehensive data on pre-admission patient characteristics, including cognitive, physical, and emotional disorders, and investigated relationships between such characteristics and long-term clinical outcomes of sepsis.

## Materials and Methods

### Study design and patients

This retrospective observational study was conducted at Saga University Hospital, in Japan. Patients aged 18 years or older who were admitted to the hospital with sepsis between August 2014 and July 2016, and whose one-year outcome was confirmed, were included in this study. Patients who developed sepsis after hospital admission or whose one-year outcome could not be confirmed were excluded from the study. All cases were categorized into two groups based on their one-year outcome: Survivors (n=31) and Deceased (n=45). The same patients were also reclassified into a group manifesting cognitive impairment (CI) and a normal, Control group.

### Variables related to patient characteristics and medical history

Patient data, including age, percentage of patients over 65 years old, sex, prior hospitalizations, transfer cases, and past medical problems were recorded, including conditions such as such as hypertension, arrhythmia, cardiovascular disease, peripheral vascular disease, cerebrovascular disease, diabetes mellitus, gastrointestinal and hepatobiliary disease, hyperlipidemia, malignancy, chronic respiratory disease, and chronic kidney disease.

### Clinical scores and blood sampling

We calculated several clinical scores as follows: the systemic inflammatory response syndrome (SIRS) score [14], the Japanese association for acute medicine (JAAM) disseminated intravascular coagulation (DIC) score [15], the acute physiology and chronic health evaluation (APACHE) II score, the sequential organ failure assessment (SOFA) score, and the quick SOFA (qSOFA) score [16]. SOFA scores were computed for each organ component, in addition to the total score. Organ failure was further defined as a score of 2 or more for a given organ. Evaluations were also conducted on the number of organ failures and on serum lactate levels.

### Sepsis management in our hospital and clinical outcomes

During the study period, sepsis was diagnosed and treated based on the Sepsis-2 definition [17]. Therefore, to compare data using the current definition of sepsis-3 [16], we also assessed sepsis occurrence according to sepsis-3 criteria. Additionally, data were collected on infection sources, pathogenic bacteria, and results of blood cultures. Sepsis treatments were performed by emergency physicians and critical care physicians at the facility. However, there is no institutional protocol for managing sepsis at this hospital. This study documents various treatments administered during each hospital stay, including antibiotics, antifungal and antiviral agents, mechanical ventilation, renal replacement therapy, anti-disseminated intravascular coagulation (DIC) therapy, catecholamines, steroids, insulin use, transfusions, and surgical interventions.

### Indicators for comprehensive functional assessment

Upon admission to and discharge from Saga University Hospital, patients or their relatives are interviewed by nurses for a comprehensive functional assessment. This assessment comprises various indices, such as “pre-admission indicators on activity of daily living and cognitive-emotional functions”, as well as “pre-admission levels of bed-ridden degree (“Netakiri” index).” The former encompasses basic difficulties with activities of daily living (ADL), such as required assistance with toileting and moving about the residence, as well as instrumental ADL difficulties, such as an inability leave the ward for examinations or to go to the store alone. Other factors contributing to the condition include lethargy, cognitive impairment such as memory problems, an inability to engage in simple conversations, and emotional disturbances such as depression. The Netakiri index classifies independence into four categories: “Rank J (independent in daily life)”, “Rank A (independent indoors, but needs support when going out)”, “Rank B (needs support even indoors)”, and “Rank C (in bed all day)”. The Barthel Index was also utilized to measure post-discharge indicators. This index consists of 10 domains, such as feeding, wheelchair-to-bed mobility, personal toilet, getting on and off the toilet, self-bathing, walking on level surfaces, stair-climbing, dressing, and bowel and bladder control.

### Variables in clinical outcomes

In this study, we collected clinical outcome data, including lengths of hospital stays, transfers to other hospitals, hospital mortality, and one-year survival rates. The composite outcome of death or tendency to be bedridden (Rank B or C of Netakiri index) at discharge was defined and evaluated for correlation with cognitive impairment and control.

### Statistical analysis

Data access for this study was initiated on February 6, 2018. Only HK, the person in charge of data analysis, had access to information that could identify individual participants after data collection. After anonymization, other authors also used these data. To evaluate factors associated with long-term outcomes, patients were categorized based on death within a year and were subjected to univariate analysis. The median (interquartile range; IQR) was used to represent continuous variables, whereas numbers (percentages) were employed for categorical/nominal variables in both groups. The Mann-Whitney U test was utilized for continuous variables, whereas the Chi-square test and Fisher’s exact test were applied to categorical variables. Statistical significance was defined as p < 0.05.

Logistic regression analysis was conducted to identify predictors of one-year survival, utilizing cognitive impairment as well as factors that showed significant differences in the univariate analysis. Several models were created, and odds ratios were calculated. Finally, we constructed Kaplan-Meier curves to analyze presence or absence of cognitive impairment and conducted log-rank tests. All statistical analyses were performed utilizing JMP® Pro 16.1.0 software (SAS Institute, Cary, NC, USA).

## Results

During the two-year observational period, our hospital diagnosed 125 patients with sepsis (Fig 1). Of these patients, 83 were initially hospitalized for sepsis, and 42 were diagnosed with sepsis after being hospitalized for another medical condition. After excluding 7 patients whose 1-year outcomes were unknown, the present study enrolled 76 patients. All participants were categorized as survivors(N=31) or deceased (N=45) 1-year after discharge.

**Fig 1.**
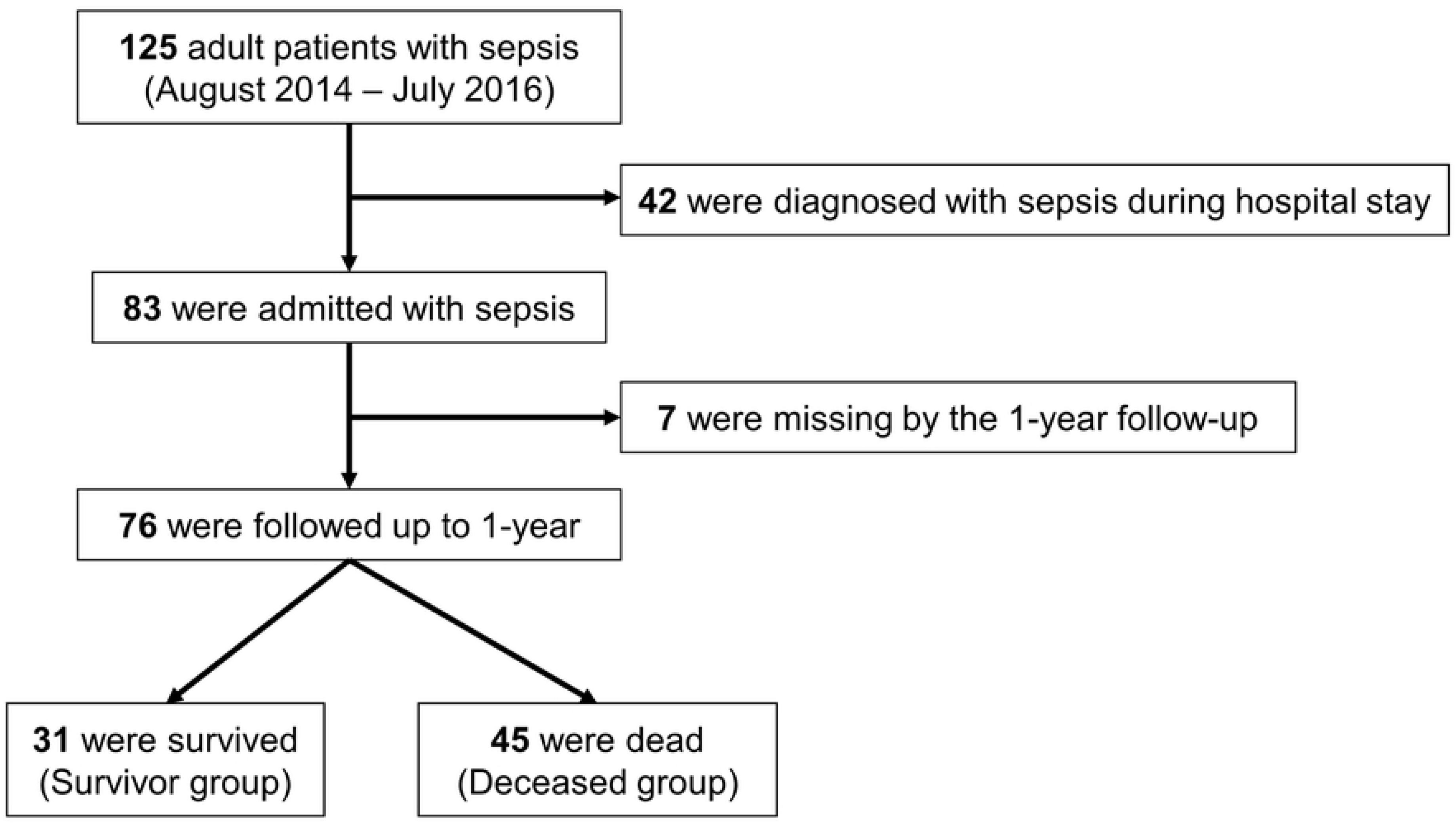
Flow diagram of the present study.

During the study period, 125 adult patients with sepsis were identified. After excluding 42 patients who were diagnosed after admission, as well as 7 patients whose 1-year outcome was unknown, 76 patients (31 survivors and 45 deceased) were enrolled.

Table 1 presents results of univariate analyses of factors related to patient characteristics and clinical scores associated with 1-year outcomes. Survivors included significantly fewer males (S: 35.5% vs. D: 80.0%, P<0.0001); however, there were no significant differences with regard to age, percentage of older patients, history of hospitalization within 1-year, or transfers to other institutions. The arrhythmia ratio was significantly lower among survivors than deceased patients in terms of past medical history (3.2 % vs. 26.7 %, P=0.01). Other medical conditions, however, were statistically equivalent in both groups. While some clinical scores, including SIRS, JAAM DIC, and qSOFA, were the same in both groups, the median values of the APACHEII score (18 vs. 22, P=0.02) and the total SOFA score (5 vs. 8, P<0.0001) were considerably lower among survivors than among deceased patients. SOFA scores for each organ of survivors were significantly lower in the central nervous system (CNS)(1 vs. 2, P=0.002), respiratory system (1 vs. 2, P=0.01), and liver (0 vs. 0, P=0.04), respectively. Additionally, the proportion of failure for each organ was significantly lower in the CNS (16.1% vs. 53.3%, P=0.0016), respiratory system (22.6% vs. 51.1%, P=0.01), and coagulation system (19.4% vs. 51.1%, P=0.005) of survivors compared to deceased patients. Consequently, the incidence of organ failure was significantly lower among survivors (1 vs. 3, P<0.0001).

**Table 1.**
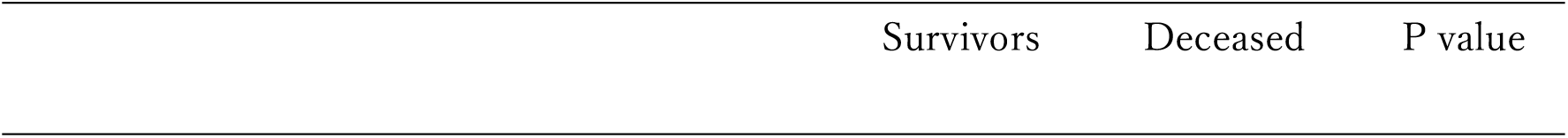
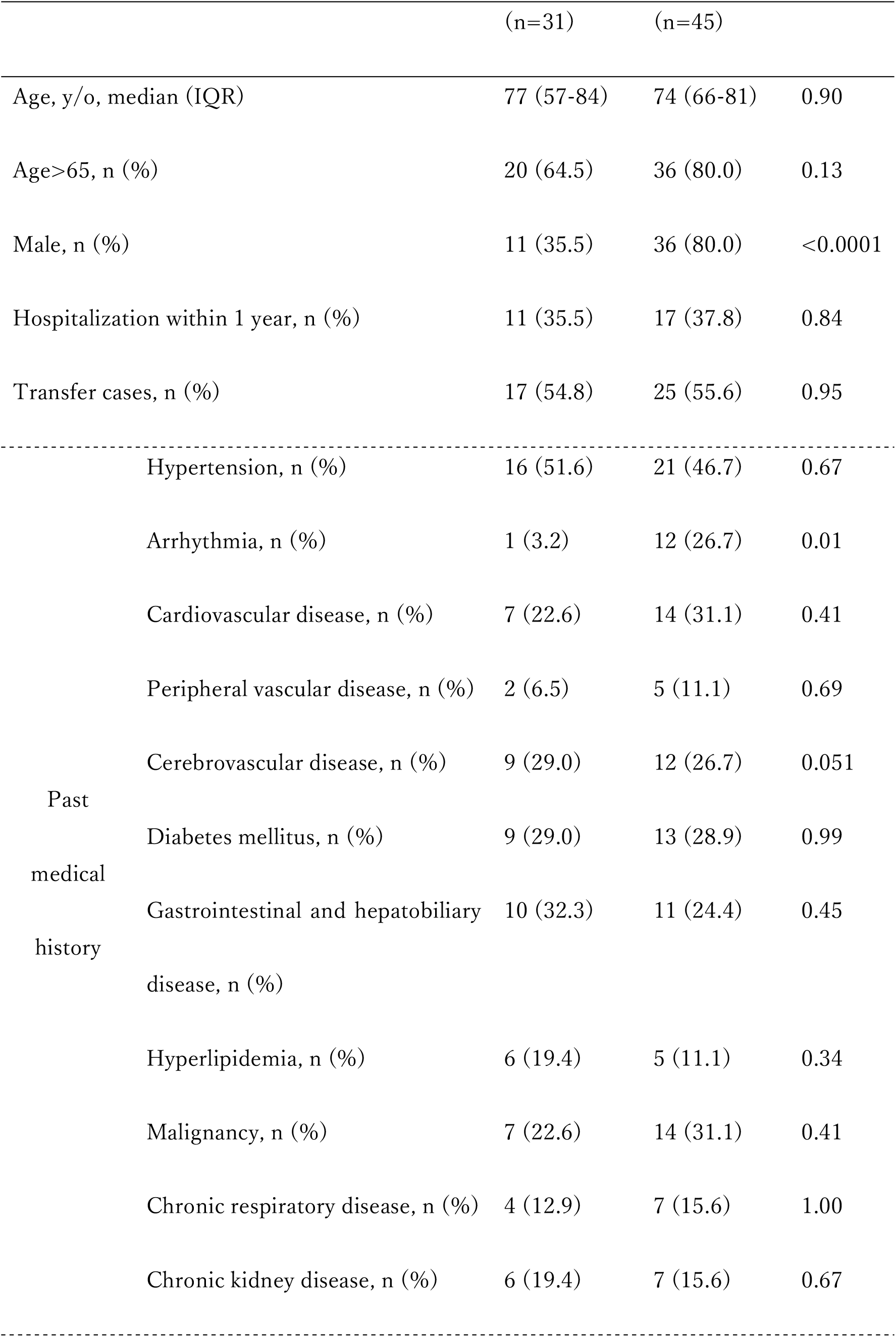
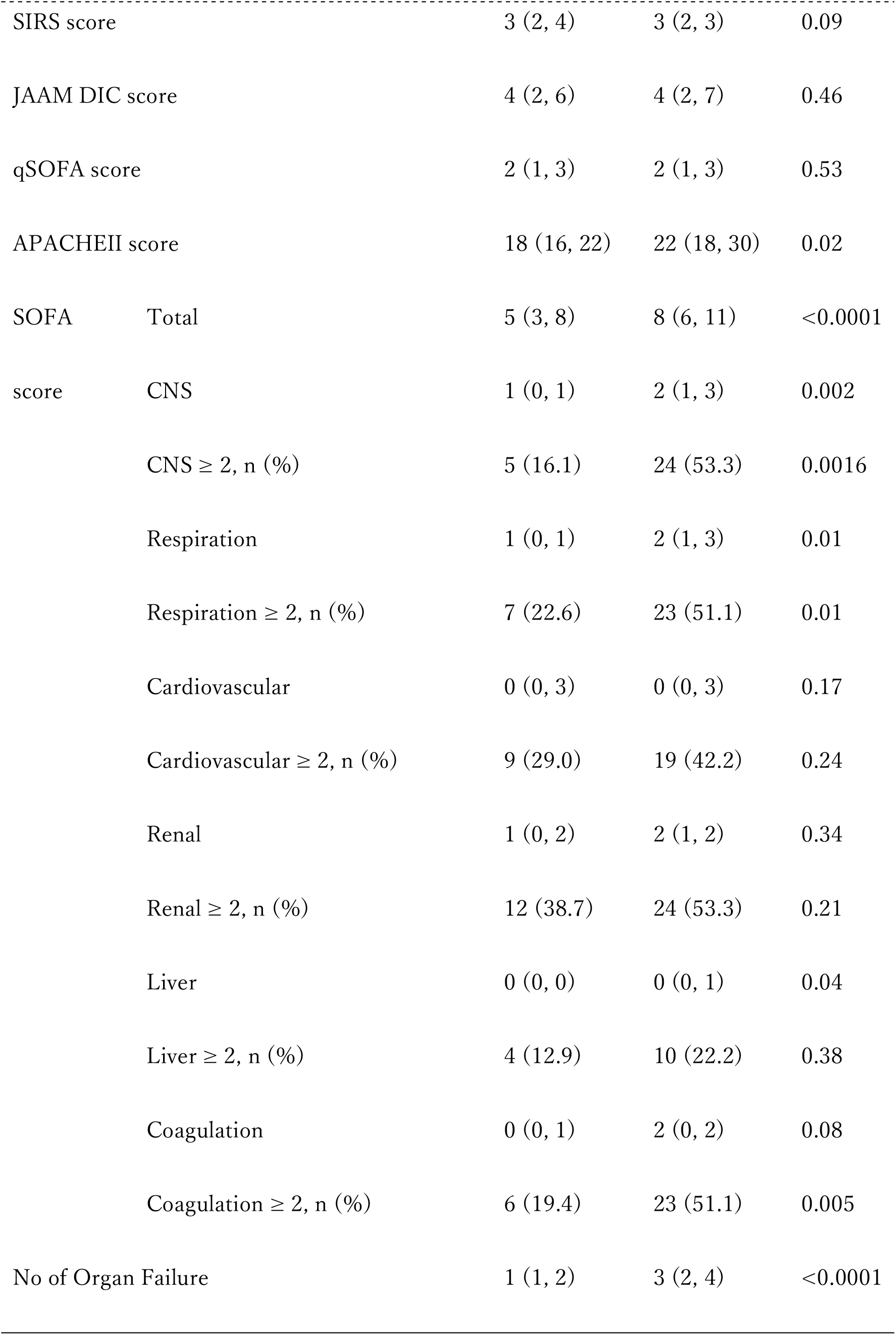
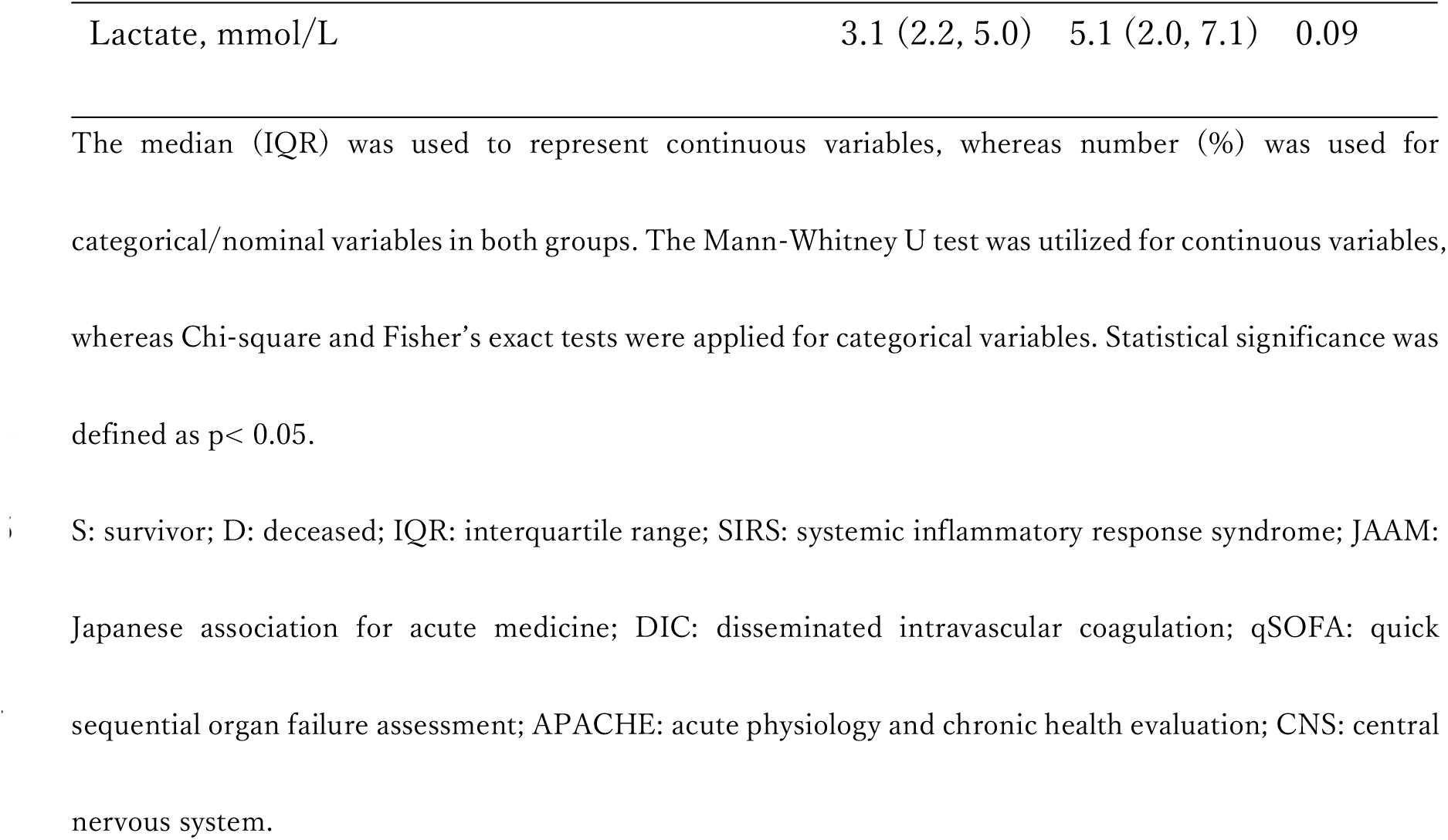
Patient characteristics and clinical scores of survivors and deceased patients.

Table 2 provides an overview of parameters related to infections. In regard to sepsis-2, survivors had significantly fewer severe cases, such as septic shock (p=0.02). Of 76 septic patients with Sepsis-2, 53 patients (69.7%) were diagnosed based on the sepsis-3 definition, with 20 survivors and 33 deceased patients diagnosed with sepsis and septic shock. A similar trend was observed using the sepsis-3-based classification; however, no significant difference was found. Furthermore, there was statistical equality among sources of infection, pathogenic bacteria, and the number of positive blood cultures.

**Table 2.** Indicators on infections and sepsis.

|  |  | S group (n=31) | NS group (n=45) | p value |
| --- | --- | --- | --- | --- |
| Sepsis-2<br>definition | Sepsis, n (%) | 11 (35.5) | 5 (11.1) | 0.02 |
|  | Severe sepsis, n (%) | 8 (25.8) | 11 (24.4) |  |
|  | Septic shock, n (%) | 12 (38.7) | 29 (64.4) |  |
| Sepsis-3<br>definition | Sepsis, n (%) | 8/20 (40.0) | 9/33 (27.3) | 0.39 |
|  | Septic shock, n (%) | 12/20 (60.0) | 24/33 (72.7) |  |
| Infection<br>sources | Respiratory, n (%) | 2 (6.5) | 13 (28.9) | 0.10 |
|  | Urinary, n (%) | 10 (32.3) | 7 (15.6) |  |
|  | Abdominal, n (%) | 9 (29.0) | 10 (22.2) |  |
|  | Soft tissue, n (%) | 4 (12.9) | 8 (17.8) |  |
|  | Others, n (%) | 6 (19.4) | 7 (15.6) |  |
| Pathogenic<br>bacteria | Gram-positive, n (%) | 13 (41.9) | 21 (46.7) | 0.47 |
|  | Gram-negative, n (%) | 12 (38.7) | 17 (37.8) |  |
|  | Mixed-type, n (%) | 1 (3.2) | 4 (8.9) |  |
|  | Not detected, n (%) | 5 (16.1) | 3 (6.7) |  |
| Positive blood culture, n (%) |  | 4 (15.4) | 9 (21.4) | 0.54 |

Number (%) was used as categorical/nominal variables in both groups. Chi-square and Fisher's exact tests
were applied for categorical variables. Statistical significance was defined as $p < 0.05$ .
S: survivor; D: deceased.

Pre-admission comprehensive functional assessment indices are shown in Table 3. Survivors had significantly lower rates of ADL difficulties (25.8% vs. 48.9%, p=0.04) and cognitive impairment (22.6% vs. 46.7%, P=0.03) compared to patients who later deceased. The two groups were statistically similar in terms of IADL difficulties, decreased motivation, emotional disturbance, and Netakiri index.

**Table 3.**
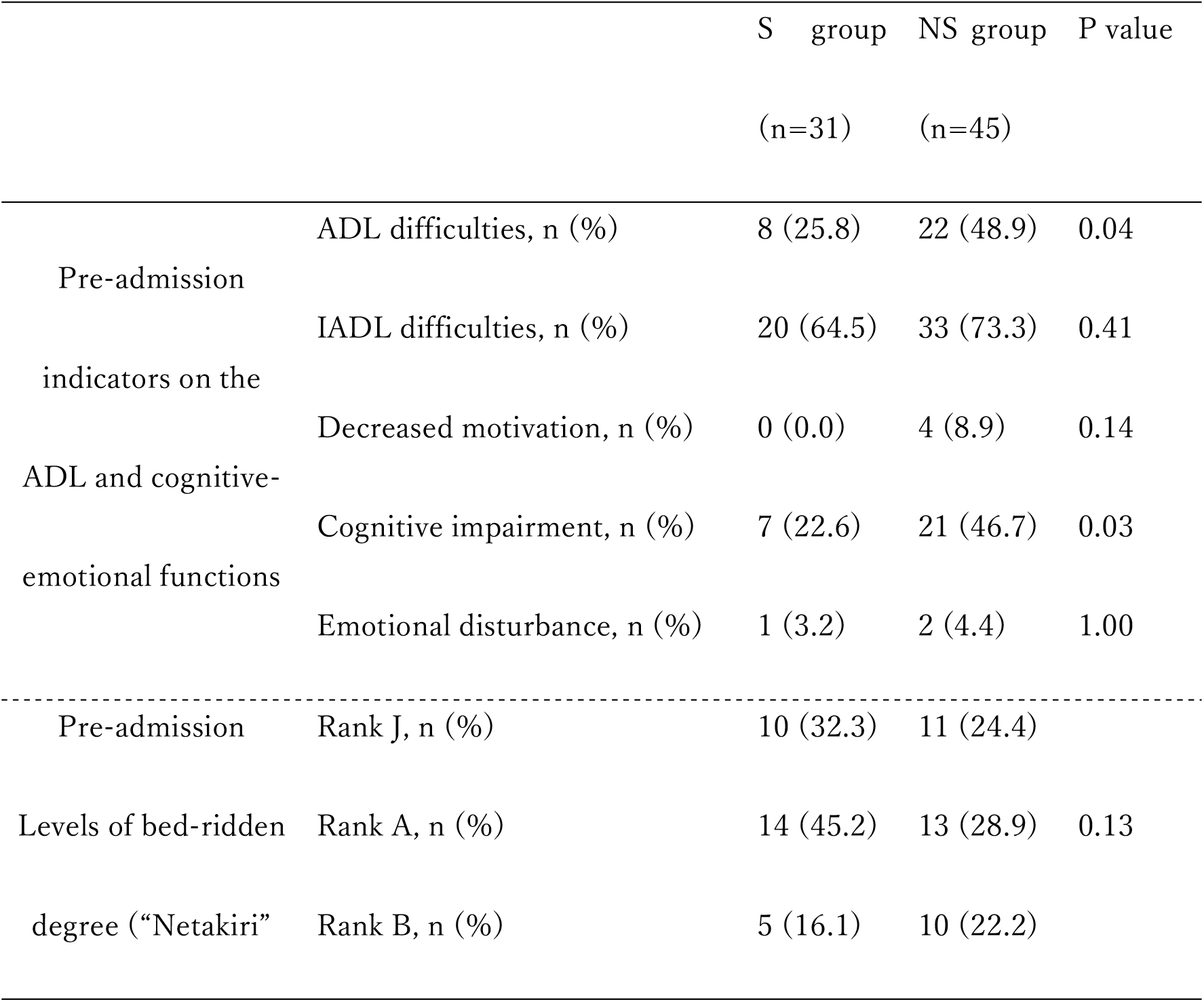
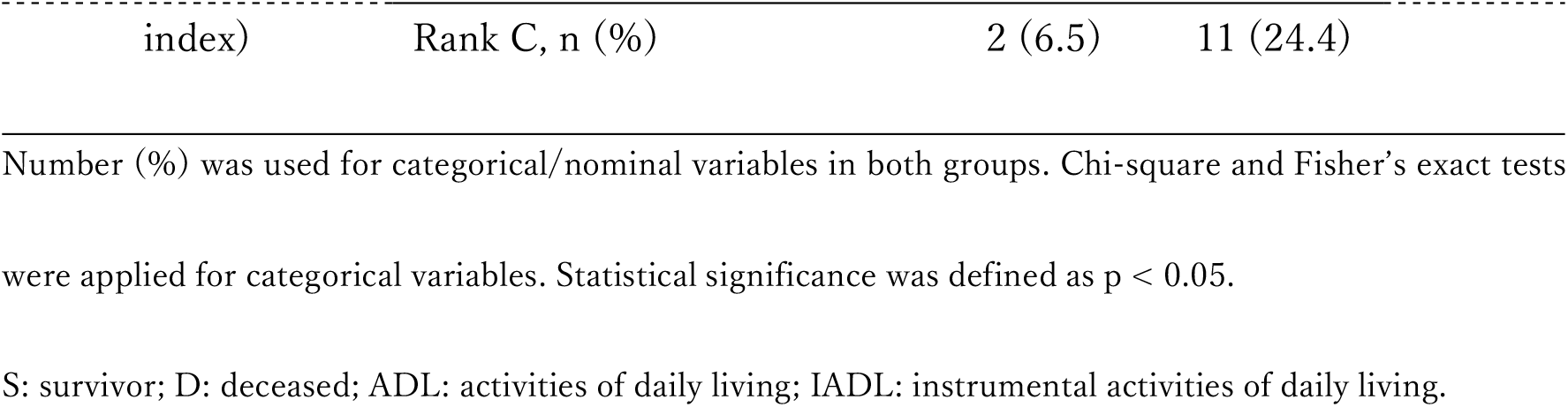
Comprehensive functional assessment indices before admission.

Table 4 provides treatment and clinical outcomes. Survivors had significantly lower rates of treatment history with catecholamines (51.6% vs. 84.4%, p=0.002) and steroids (25.8% vs. 51.1%, p=0.03) than patients who later deceased. No significant differences were found in other treatments irrespective of 1-year outcome. Survivors showed significant improvements in clinical outcomes compared to the control group, with longer hospital stays (28 days vs. 12 days, p=0.0006), lower hospital mortality (0.0% vs. 62.2%, P<0.0001), and longer survival time within one year (365 days vs. 15 days, p<0.0001).

**Table 4.** Treatments and clinical outcomes.

|  | S group (n=31) | NS group (n=45) | P value |
| --- | --- | --- | --- |
| Antibiotics, n (%) | 31 (100.0) | 45 (100.0) | --- |
| Antifungal agents, n (%) | 3 (9.7) | 6 (13.3) | 0.73 |
| Antiviral agents, n (%) | 1 (3.2) | 1 (2.2) | 1.00 |
| Mechanical ventilation, n (%) | 11 (35.5) | 26 (57.8) | 0.06 |
| Renal replacement therapy, n (%) | 15 (48.4) | 16 (35.6) | 0.26 |
| Anti-DIC agents, n (%) | 18 (58.1) | 26 (57.8) | 0.98 |
| Catecholamines, n (%) | 16 (51.6) | 38 (84.4) | 0.002 |
| Steroids, n (%) | 8 (25.8) | 23 (51.1) | 0.03 |
| Insulin use, n (%) | 8 (25.8) | 11 (24.4) | 0.89 |
| Transfusions, n (%) | 5 (16.1) | 12 (26.7) | 0.40 |
| Surgical interventions, n (%) | 12 (38.7) | 13 (28.9) | 0.37 |
| Length of hospital stay, days | 28 (15, 69) | 12 (3, 24) | 0.0006 |
| Hospital mortality, n (%) | 0 (0.0) | 28 (62.2) | <0.0001 |
| Transferred to the other hospital, n (%) | 19 (65.5) | 14 (82.4) | 0.22 |
| Survival days within 1 year, days | 365 (365, 365) | 15 (3, 77) | <0.0001 |

Median (IQR) was used to represent continuous variables, whereas number (%) was used for categorical/nominal variables in both groups. The Mann-Whitney U test was utilized for continuous variables, whereas Chi-square and Fisher's exact tests were applied for categorical variables. Statistical significance was defined as $p < 0.05$ .
S: survivor; D: deceased; IQR: interquartile range; DIC: disseminated intravascular coagulation.

Next, we focused on cognitive impairment and developed several logistic regression models using factors that demonstrated significant differences through univariate analysis (Table 5). Remarkably, cognitive impairment was an independent predictor of worse 1-year survival for all models. For example, in model 3, cognitive impairment was an independent predictor of a poor outcome after one year (odds ratio 0.184, 95% confidence interval 0.047- 0.722, p=0.0152).

**Table 5.**
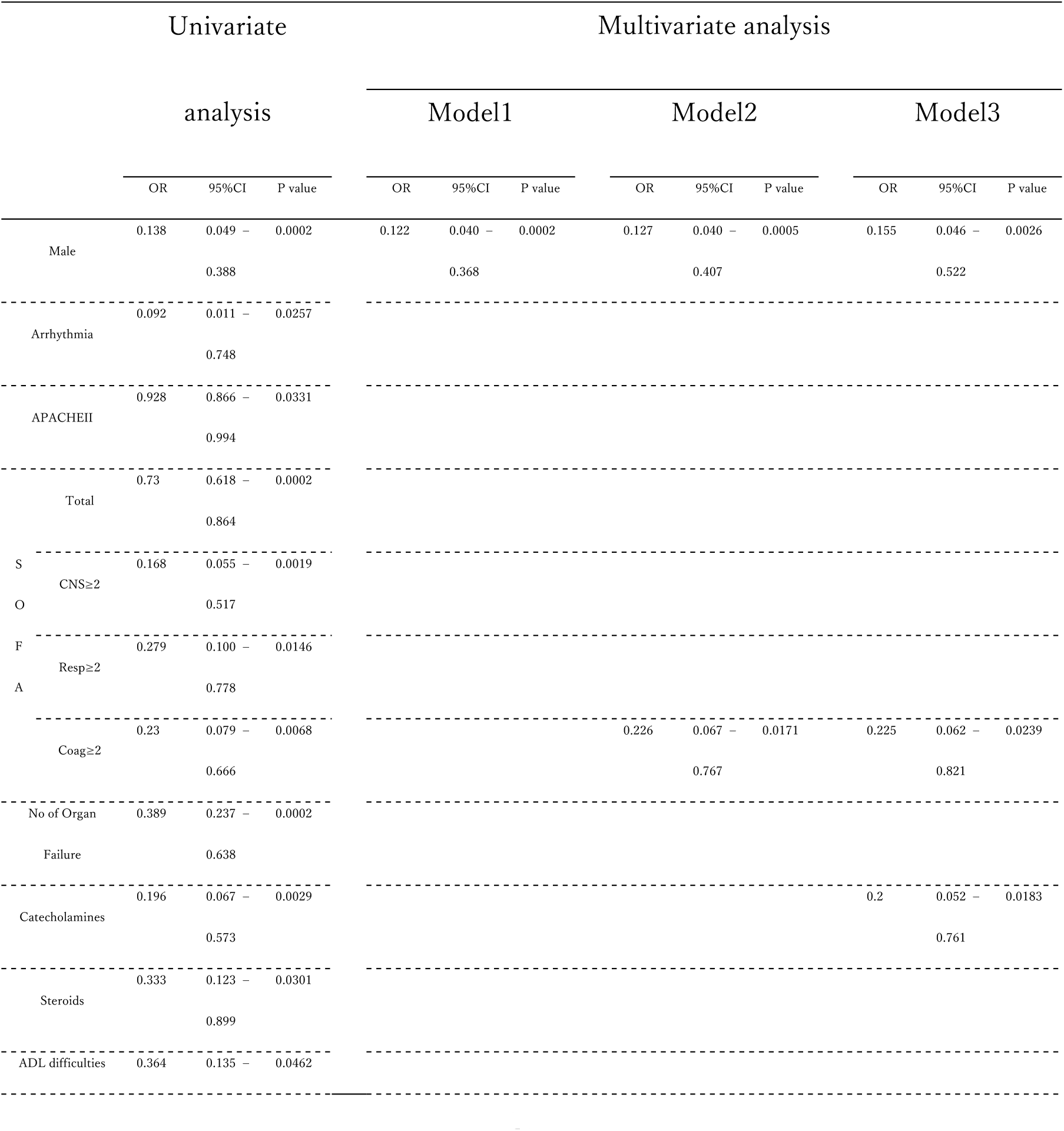
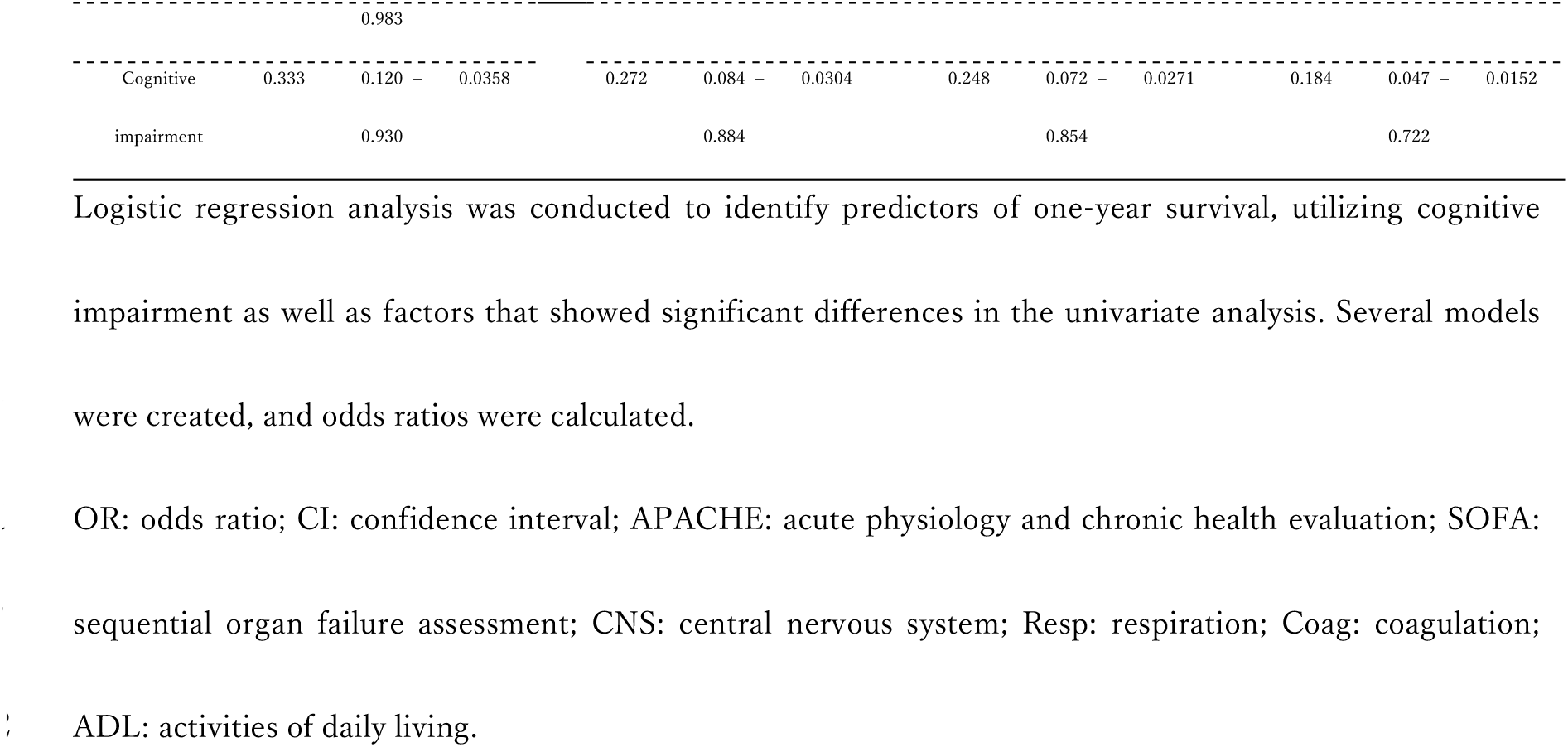
Univariate and multivariate analyses for predicting 1-year survival.

Based on results of multivariate analysis, we conducted a univariate analysis of the cognitive impairment (CI) and Control groups (Table 6). The CI group exhibited a significantly higher proportion of patients over 65 years of age (CI group; 89.3% vs. Control group; 64.6%, P=0.03) and higher APACHEII scores (23 vs. 19, p=0.008) than the control group. Moreover, there were fewer patients who received renal replacement therapy (21.4% vs. 52.1%, p=0.009) or anti-DIC therapy (42.9% vs. 66.7%, p=0.04). Surprisingly, the 1-year survival rate of the CI group was significantly lower than that of the Control group (25.0% vs. 50.0%, p=0.03). Cognitive impairment was linked to greater difficulties with ADL (64.3% vs. 25.0%, p=0.0007) and IADL (96.4% vs. 54.2%, p<0.0001) before hospitalization and a significantly lower Barthel index upon discharge (0 vs. 45, p=0.0001). Furthermore, cognitive impairment was significantly related to being bedridden, as indicated by the Netakiri index, both before admission and upon discharge. The CI group had a significantly higher occurrence of the composite outcome, consisting of death at discharge or bedridden status at discharge, compared to the Control group (100.0% vs. 75.0%, p=0.0028). The log-rank test revealed a significantly lower survival rate in the CI group as opposed to the Control group (p=0.0332) (Fig 2).

**Fig 2.**
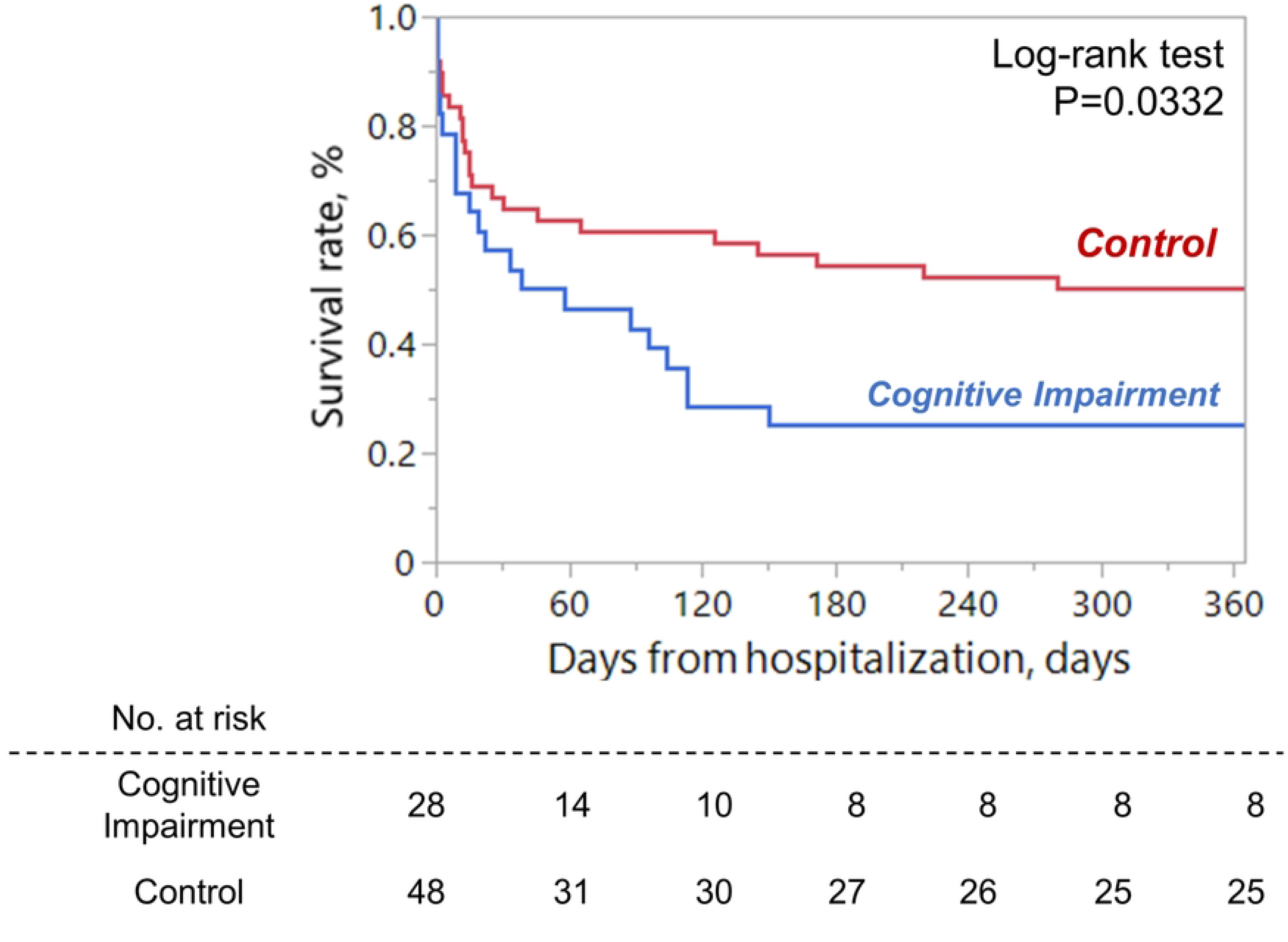
Cognitive impairment and 1-year mortality.

**Table 6.**
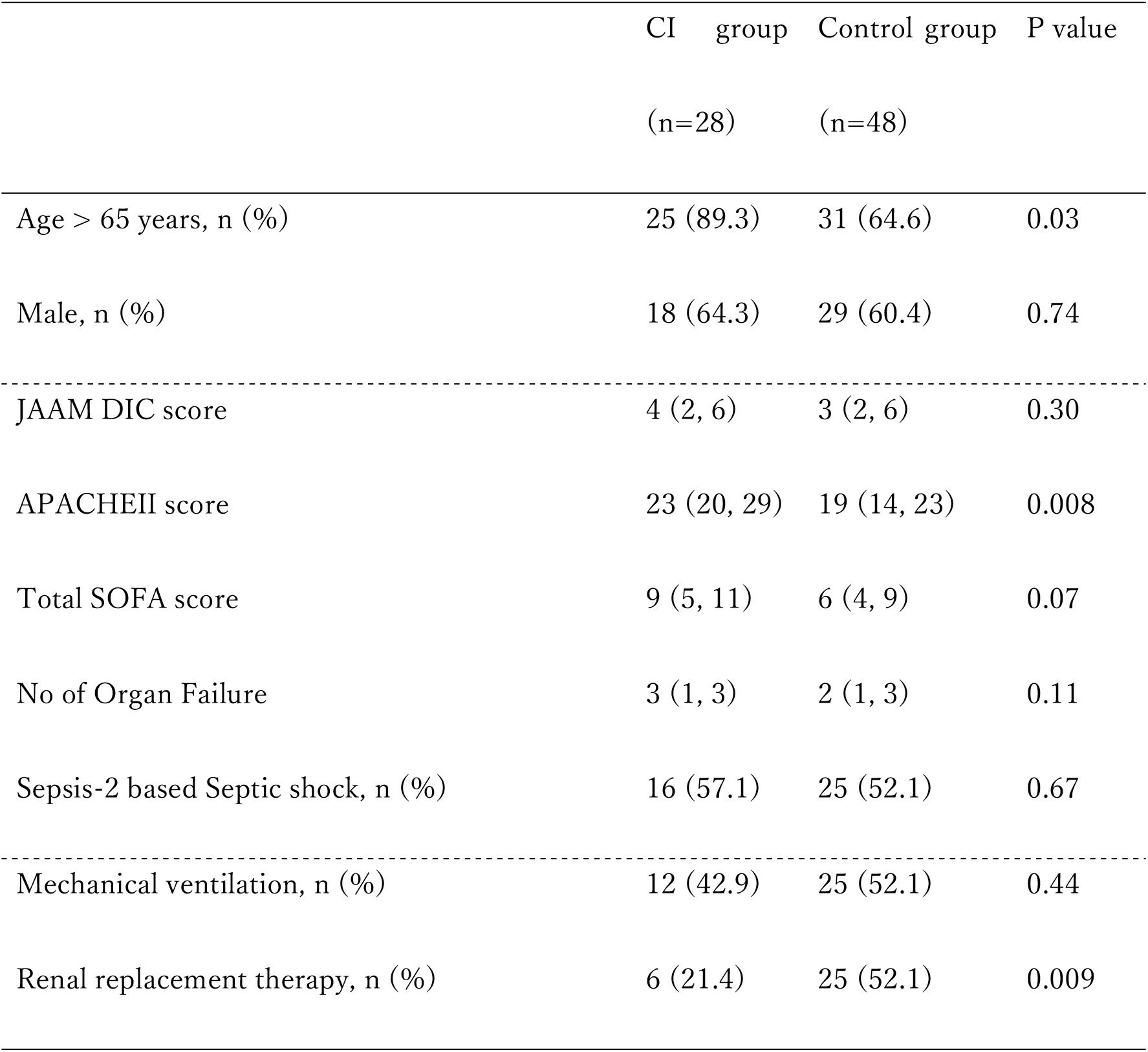
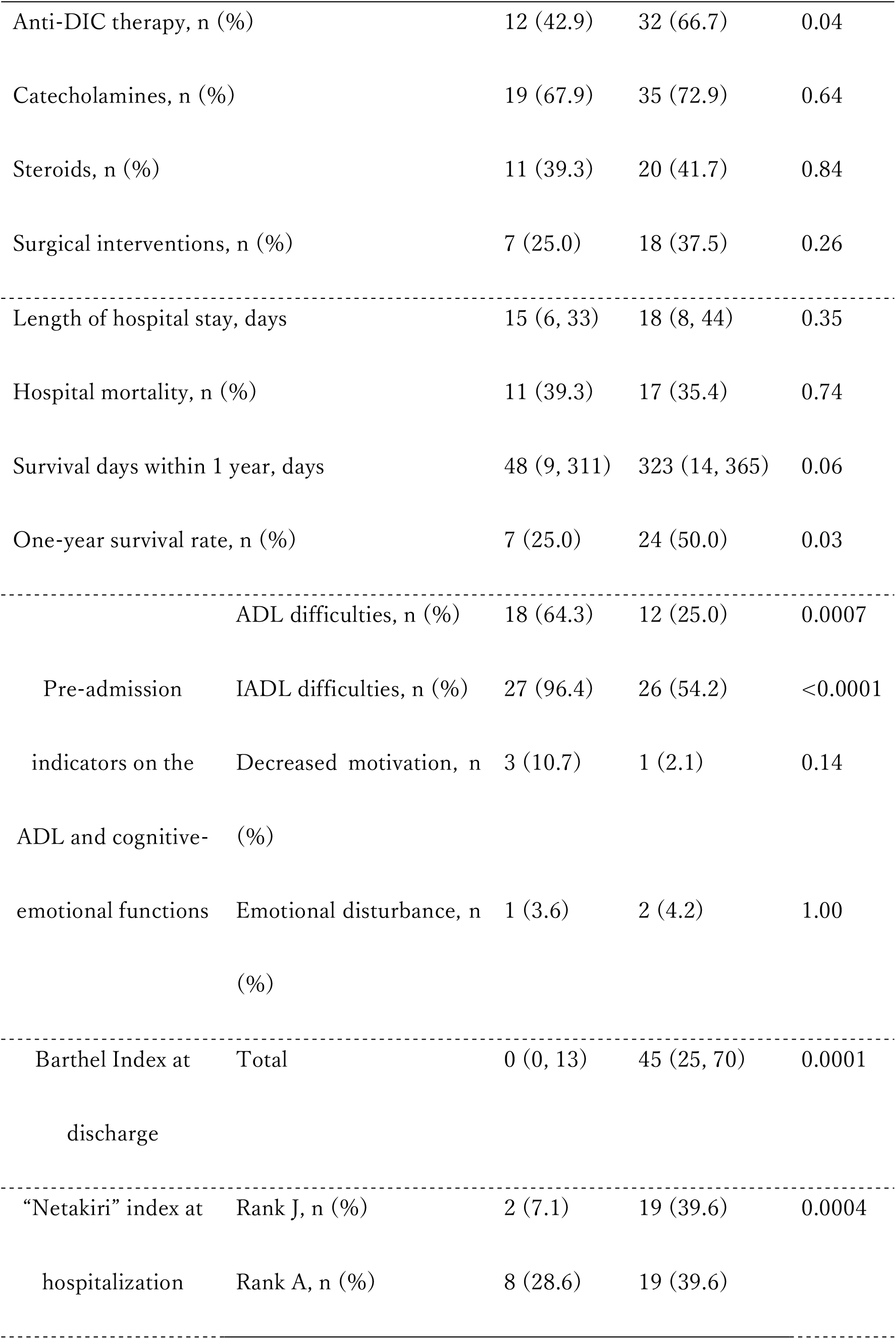
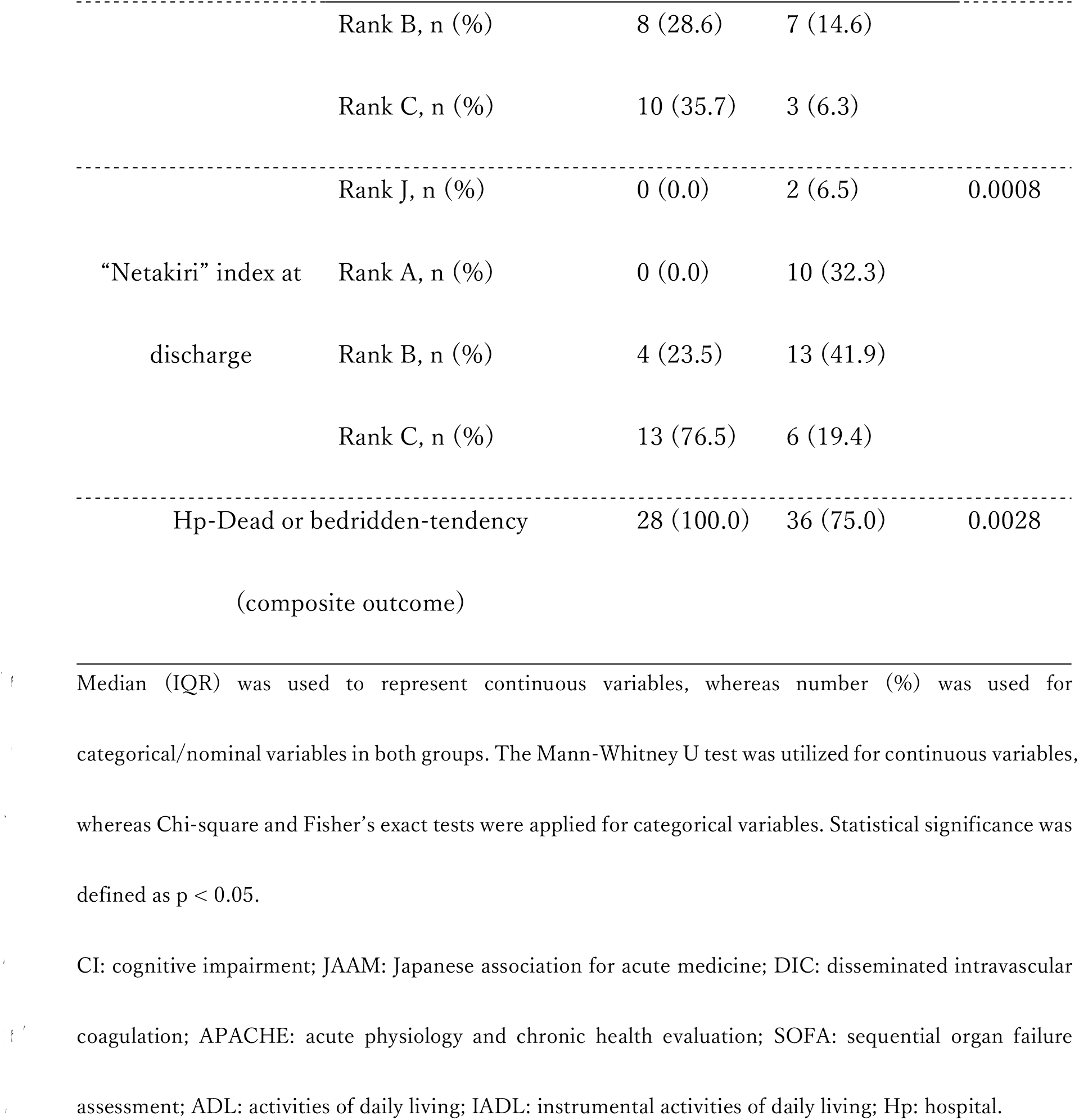
Univariate analysis for cognitive impairment between the CI and Control groups.

Survival analysis with the log-rank test demonstrated that the cognitive impairment group had significantly lower survival.

## Discussion

This retrospective study revealed that within a year of hospitalization and treatment for sepsis, 59% of patients died. Cognitive impairment was identified as an independent predictor of unfavorable outcomes one year after sepsis. Sepsis patients with cognitive impairment were more likely to be elderly, had greater clinical severity, and were also more likely to be bedridden and physically impaired. Even if these patients could have been saved, they were all left bedridden at discharge, and their one-year survival rate was only 25%.

In this study, 28 patients (36.8%) died during hospitalization, with a median hospital stay of just 12 days, despite receiving intensive sepsis care. Our patient cohort exhibited greater illness severity than the reported Japanese sepsis patient population, with a significantly higher rate of organ support and about twice the mortality rate [18]. It is noteworthy that 17 (35.4%) of the 48 patients who were discharged alive from our hospital died within a year of discharge. This suggests that mortality rates remain high throughout the first-year post-sepsis onset, a similar level to that of the acute phase. Wang and colleagues reported an adjusted hazard ratio of 13.07 for sepsis-related death in the first year following a sepsis event, compared to those who had no sepsis [8]. This ratio remains 2-fold higher as much as five years post-event. Moreover, a systematic review and meta-analysis of epidemiological studies revealed a 1-year post-acute mortality rate of 16.1% for sepsis, although these studies showed a high degree of heterogeneity, and a consensus has yet to be reached [6].

When interpreting clinical outcomes of sepsis, it is crucial to consider the diverse patient characteristics associated with sepsis. In the present retrospective study, 56 (73.7%) of the 76 patients enrolled were elderly, and 60 (78.9%) were diagnosed with severe sepsis and septic shock, based on the sepsis-2 definition. Additionally, there were fewer cases of respiratory infections as sources of sepsis, but more cases of urinary tract and soft tissue infections compared to recent reports from Japan [18, 19]. As we expected, sepsis severity was associated with death within 1-year; however, it was surprising that infection sources, bacterial profiles, and blood culture positivity were not related to long-term outcomes. This suggests that factors other than sepsis severity may significantly affect long-term prognosis following sepsis. Post-intensive care syndrome (PICS) has recently received considerable attention and could be a prominent example of such factors. A single-center prospective study conducted in Japan found that 70% of patients experienced PICS three months after contracting sepsis [20]. The study identified Glasgow coma scale (GCS) on day 7 as an independent, reliable marker for PICS, rather than any variables related to sepsis. We confirmed the potential impact of various multidisciplinary treatments, including administration of catecholamines and steroids, as well as ventilatory management, on long-term prognosis. However, we did not evaluate PICS in our investigation, but we plan to explore further evidence in future studies.

The most significant finding of this study is that cognitive impairment prior to admission is an independent predictor of 1-year mortality from sepsis. Little evidence was available regarding the prevalence of cognitive impairment at the time of sepsis [10]. In a prospective cohort study, Iwashyna et al. reported that 6.1% of patients who survived severe sepsis had cognitive impairment before disease onset [10]. Upon admission to our emergency department, 36.8% of patients enrolled in this study had cognitive impairment, which was strongly associated with clinical outcome in sepsis, in contrast to previous reports. Sepsis patients with cognitive impairment were not only older and clinically more severely ill, but they were also significantly associated with poorer ADL and IADL. They tended to be bedridden and had a lower Barthel indices at discharge. Interestingly, all septic patients with cognitive impairment were also bedridden or deceased upon discharge. These results indicate that cognitive impairment in sepsis patients increases the severity of both short-term and long-term outcomes.

Furthermore, the issue of cognitive impairment after sepsis is a significant concern for ICU patients. Iwasyna et al. reported that 16.7% of sepsis survivors had moderate to severe cognitive impairment after severe sepsis [10]. A recent systematic review found that cognitive impairment of sepsis survivors ranged from 12.5 to 21% [11]. Pathophysiological changes, such as blood-brain barrier dysregulation, neuroinflammation, neurotransmitter dysfunction, and neuronal loss are thought to be responsible for long-term cognitive impairments [21].

More recently, chronic conditions of sepsis-associated encephalopathy have been recognized, including memory impairment, attention deficit, language impairment, and executive dysfunction, which are present in more than half of sepsis survivors at the time of hospital discharge [22].

In light of the above, early confirmation as well as post-sepsis follow-up may help to improve long-term outcomes in sepsis patients. When patients with sepsis arrive at the emergency department, it is crucial to assess not only their medical history, medications, family history, and social problems, but also their physical limitations, cognitive impairment, and emotional distress, especially in elderly patients. At the very least, our data suggest that sepsis patients with cognitive impairment are highly prone to be bedridden or dead at hospital discharge. Early rehabilitation is one of the few effective options to prevent patients from becoming bedridden. Recent guidelines on rehabilitation for PICS indicate that cognitive rehabilitation can prevent the onset of PICS and can reduce symptoms such as delirium, attention deficit, memory impairment, executive dysfunction, and spatial cognitive impairment [23]. Additionally, seamless cooperation between inpatient and outpatient rehabilitation programs should not be overlooked. In the present study, the median length of hospital stay was only 16 days, and sepsis patients stay in smaller community hospitals, nursing facilities, or at home for more than 300 days. In the future, it is important for communities to consider consistent rehabilitation for sepsis patients after discharge from the hospital.

This study has several limitations. Its small sample size, single-center status, and retrospective nature limit generalizability. In addition, most enrolled sepsis patients were hospitalized based on the sepsis-2 definition. Therefore, results of this study may not be applicable to sepsis-3 patients. All assessments of patient physical difficulties, cognitive impairment, emotional problems, and the Netakiri index were performed by nurses without officially certified assessment tools. This shows that there are limits to the objectivity of assessors.

## Conclusions

This retrospective study found that 59% of sepsis patients died within 1-year of release from the hospital. Cognitive impairment was the most reliable independent predictor of poor outcomes. Sepsis patients with cognitive impairment are likely to be bedridden or dead at hospital discharge. Future research is warranted regarding early detection of cognitive impairment and effectiveness of rehabilitation programs.

## Acknowledgments

We thank Steven D. Aird (https://www.sda-technical-editor.org) for editing the manuscript.

## Author contributions

Conceptualization: Hiroyuki Koami, Yutaro Furukawa, Yuichiro Sakamoto. Data curation: Hiroyuki Koami.

Formal Analysis: Hiroyuki Koami. Investigation: Hiroyuki Koami.

Methodology: Hiroyuki Koami.

Project Administration: Hiroyuki Koami, Yuichiro Sakamoto. Supervision: Yuichiro Sakamoto.

Validation: Hiroyuki Koami, Ayaka Matsuoka, Kota Shinada. Visualization: Hiroyuki Koami.

Writing-Original Draft Preparation: Hiroyuki Koami.

Writing-Review & Editing: Yutaro Furukawa, Kosuke Mouri, Ayaka Matsuoka, Kota Shinada, Kento Nakayama, Sachiko Iwanaga, Shogo Narumi, Mayuko Koba, Yuichiro Sakamoto.

## Data Availability Statement

Datasets generated and/or analyzed during the present study are not publicly available due to ethical restrictions associated with personal information, but anonymized data are available from the corresponding author on request.

## Disclosure

Approval of the research protocol: This study was conducted in accordance with the declaration of Helsinki (Fortaleza, Brazil, October 2013) and was approved by the ethics committee of Saga University Hospital (protocol identification number; 2017-11-R-11).

Informed consent: The ethics committee waived the requirement for informed consent due to the retrospective nature of the study.

## References

1. Fleischmann C, Scherag A, Adhikari NK, Hartog CS, Tsaganos T, Schlattmann P, et al. Assessment of Global Incidence and Mortality of Hospital-treated Sepsis. Current Estimates and Limitations. American journal of respiratory and critical care medicine. 2016;193(3):259–72. doi: 10.1164/rccm.201504-0781OC. PubMed PMID: 26414292.

2. Dellinger RP, Carlet JM, Masur H, Gerlach H, Calandra T, Cohen J, et al. Surviving Sepsis Campaign guidelines for management of severe sepsis and septic shock. Intensive Care Med. 2004;30(4):536–55. doi: 10.1007/s00134-004-2210-z. PubMed PMID: 14997291.

3. Angus DC, Carlet J, Brussels Roundtable P. Surviving intensive care: a report from the 2002 Brussels Roundtable. Intensive Care Med. 2003;29(3):368–77. Epub 20030121. doi: 10.1007/s00134-002-1624-8. PubMed PMID: 12536269.

4. Maley JH, Mikkelsen ME. Short-term Gains with Long-term Consequences: The Evolving Story of Sepsis Survivorship. Clinics in chest medicine. 2016;37(2):367–80. Epub 20160310. doi: 10.1016/j.ccm.2016.01.017. PubMed PMID: 27229651.

5. Prescott HC, Angus DC. Enhancing Recovery From Sepsis: A Review. JAMA : the journal of the American Medical Association. 2018;319(1):62–75. doi: 10.1001/jama.2017.17687. PubMed PMID: 29297082; PubMed Central PMCID: PMCPMC5839473.

6. Shankar-Hari M, Ambler M, Mahalingasivam V, Jones A, Rowan K, Rubenfeld GD. Evidence for a causal link between sepsis and long-term mortality: a systematic review of epidemiologic studies. Critical care. 2016;20:101. Epub 20160413. doi: 10.1186/s13054-016-1276-7. PubMed PMID: 27075205; PubMed Central PMCID: PMCPMC4831092.

7. Winters BD, Eberlein M, Leung J, Needham DM, Pronovost PJ, Sevransky JE. Long- term mortality and quality of life in sepsis: a systematic review. Crit Care Med. 2010;38(5):1276–83. doi: 10.1097/CCM.0b013e3181d8cc1d. PubMed PMID: 20308885.

8. Wang HE, Szychowski JM, Griffin R, Safford MM, Shapiro NI, Howard G. Long-term mortality after community-acquired sepsis: a longitudinal population-based cohort study. BMJ open. 2014;4(1):e004283. Epub 20140117. doi: 10.1136/bmjopen-2013-004283. PubMed PMID: 24441058; PubMed Central PMCID: PMCPMC3902401.

9. Guerra C, Hua M, Wunsch H. Risk of a Diagnosis of Dementia for Elderly Medicare Beneficiaries after Intensive Care. Anesthesiology. 2015;123(5):1105–12. doi: 10.1097/ALN.0000000000000821. PubMed PMID: 26270938; PubMed Central PMCID: PMCPMC4618156.

10. Iwashyna TJ, Ely EW, Smith DM, Langa KM. Long-term cognitive impairment and functional disability among survivors of severe sepsis. JAMA : the journal of the American Medical Association. 2010;304(16):1787–94. doi: 10.1001/jama.2010.1553. PubMed PMID: 20978258; PubMed Central PMCID: PMCPMC3345288.

11. Calsavara AJC, Nobre V, Barichello T, Teixeira AL. Post-sepsis cognitive impairment and associated risk factors: A systematic review. Australian critical care : official journal of the Confederation of Australian Critical Care Nurses. 2018;31(4):242–53. Epub 20170620. doi: 10.1016/j.aucc.2017.06.001. PubMed PMID: 28645546.

12. Needham DM, Colantuoni E, Dinglas VD, Hough CL, Wozniak AW, Jackson JC, et al. Rosuvastatin versus placebo for delirium in intensive care and subsequent cognitive impairment in patients with sepsis-associated acute respiratory distress syndrome: an ancillary study to a randomised controlled trial. Lancet Respir Med. 2016;4(3):203–12. Epub 20160129. doi: 10.1016/S2213-2600(16)00005-9. PubMed PMID: 26832963; PubMed Central PMCID: PMCPMC4792772.

13. Ehlenbach WJ, Gilmore-Bykovskyi A, Repplinger MD, Westergaard RP, Jacobs EA, Kind AJH, et al. Sepsis Survivors Admitted to Skilled Nursing Facilities: Cognitive Impairment, Activities of Daily Living Dependence, and Survival. Crit Care Med. 2018;46(1):37–44. doi: 10.1097/CCM.0000000000002755. PubMed PMID: 28991827; PubMed Central PMCID: PMCPMC5858875.

14. Iba T, Nisio MD, Levy JH, Kitamura N, Thachil J. New criteria for sepsis-induced coagulopathy (SIC) following the revised sepsis definition: a retrospective analysis of a nationwide survey. BMJ open. 2017;7(9):e017046. Epub 2017/10/01. doi: 10.1136/bmjopen-2017-017046. PubMed PMID: 28963294; PubMed Central PMCID: PMCPMC5623518.

15. Gando S, Iba T, Eguchi Y, Ohtomo Y, Okamoto K, Koseki K, et al. A multicenter, prospective validation of disseminated intravascular coagulation diagnostic criteria for critically ill patients: comparing current criteria. Crit Care Med. 2006;34(3):625–31. PubMed PMID: 16521260.

16. Singer M, Deutschman CS, Seymour CW, Shankar-Hari M, Annane D, Bauer M, et al. The Third International Consensus Definitions for Sepsis and Septic Shock (Sepsis-3). JAMA : the journal of the American Medical Association. 2016;315(8):801–10. doi: 10.1001/jama.2016.0287. PubMed PMID: 26903338.

17. Levy MM, Fink MP, Marshall JC, Abraham E, Angus D, Cook D, et al. 2001 SCCM/ESICM/ACCP/ATS/SIS International Sepsis Definitions Conference. Crit Care Med. 2003;31(4):1250–6. doi: 10.1097/01.CCM.0000050454.01978.3B. PubMed PMID: 12682500.

18. Imaeda T, Oami T, Takahashi N, Saito D, Higashi A, Nakada TA. Epidemiology of sepsis in a Japanese administrative database. Acute Med Surg. 2023;10(1):e890. Epub 20231012. doi: 10.1002/ams2.890. PubMed PMID: 37841963; PubMed Central PMCID: PMCPMC10570497.

19. Wada T, Yamakawa K, Kabata D, Abe T, Ogura H, Shiraishi A, et al. Age-related differences in the survival benefit of the administration of antithrombin, recombinant human thrombomodulin, or their combination in sepsis. Scientific reports. 2022;12(1):9304. Epub 20220603. doi: 10.1038/s41598-022-13346-3. PubMed PMID: 35660774; PubMed Central PMCID: PMCPMC9166729.

20. Inoue S, Nakanishi N, Sugiyama J, Moriyama N, Miyazaki Y, Sugimoto T, et al. Prevalence and Long-Term Prognosis of Post-Intensive Care Syndrome after Sepsis: A Single- Center Prospective Observational Study. J Clin Med. 2022;11(18). Epub 20220906. doi: 10.3390/jcm11185257. PubMed PMID: 36142904; PubMed Central PMCID: PMCPMC9505847.

21. Li Y, Ji M, Yang J. Current Understanding of Long-Term Cognitive Impairment After Sepsis. Front Immunol. 2022;13:855006. Epub 20220506. doi: 10.3389/fimmu.2022.855006. PubMed PMID: 35603184; PubMed Central PMCID: PMCPMC9120941.

22. Chung HY, Wickel J, Brunkhorst FM, Geis C. Sepsis-Associated Encephalopathy: From Delirium to Dementia? J Clin Med. 2020;9(3). Epub 20200305. doi: 10.3390/jcm9030703. PubMed PMID: 32150970; PubMed Central PMCID: PMCPMC7141293.

23. Renner C, Jeitziner MM, Albert M, Brinkmann S, Diserens K, Dzialowski I, et al. Guideline on multimodal rehabilitation for patients with post-intensive care syndrome. Critical care. 2023;27(1):301. Epub 20230731. doi: 10.1186/s13054-023-04569-5. PubMed PMID: 37525219; PubMed Central PMCID: PMCPMC10392009.

